# Bioinformatical analysis identified significant genes in the PI3K-Akt signaling pathway relating with poor prognosis of skin cutaneous melanoma

**DOI:** 10.1101/2020.09.08.20190280

**Authors:** Yingzi Zhang, Yunlan Zhang, Xuyun Hu

## Abstract

Skin cutaneous melanoma (SKCM) is the most serious form of skin cancer and the reliable genetic prognostic biomarkers are not clear. The purpose of the present academic work was to identify reliable prognostic biomarkers and understand the underlying mechanism. Gene expression profiles of GSE31909, GSE104849 and GSE111766 were available from Gene Expression Omnibus (GEO). DEGs were collected from 13 melanoma samples and 12 melanocyte samples which were in the three profile datasets by GEO2R analysis. Gene ontology (GO) analysis and KEGG analysis were performed with the Database for Annotation, Visualization and Integrated Discovery (DAVID).We further established protein-protein interaction (PPI) and identify core genes with Search Tool for the Retrieval of Interacting Genes (STRING) and Molecular Complex Detection (MCODE) app in Cytoscape software. There were total of 63 consistently expressed genes in the three datasets, including eight up-regulated genes enriched in biological processes like osteoblast differentiation and extracellular matrix organization, and cell component like extracellular space, while 55 down-regulated genes enriched in positive regulation of gene expression, intracellular signal transduction, cell adhesion and apoptotic process. With prognostic information and expression data from UALCAN and Gene Expression Profiling Interactive Analysis (GEPIA), we identified three significant genes (*TNC, SPP1* and *KIT*). After Kyoto Encyclopedia of Genes and Genomes (KEGG) analysis, all three genes were located in the PI3K-Akt Signaling Pathway. Our results provide additional genetic biomarkers for SKCM patients.

## Introduction

Skin cutaneous melanoma (SKCM) is the most serious form of skin cancer. It is the fifth most common cancer and is increasing faster than any other potentially preventable cancer in the United States (1, 2). Five-year survival rates for people with melanoma depend on the stage of the disease (from stage 0 to stage Ⅳ) at the time of diagnosis. Most people with stage 0 or I melanoma can expect prolonged disease-free survival after treatment, whereas patients with more advanced stage II to IV melanoma are more likely to develop metastatic disease (2, 3). Therefore, identifying reliable prognostic biomarkers will help to predict the treatment effect and understand the underlying mechanism.

Gene expression profiles varied in different pathophysiological process. With the help of expression beadchips, we can identify differentially expressed genes (DEGs) in genome wide easily. Recent studies on SKCM produced public bioinformatical data which could be used to further identify prognostic biomarker genes (4–7). In this study, we chose microarrays of GSE31909, GSE104849 and GSE111766 from Gene Expression Omnibus (GEO, http://www.ncbi.nlm.nih.gov/geo/) (8). After GEO2R analysis, 63 common DEGs from the three datasets were collected for further study. We further established protein-protein interaction (PPI) and identified core genes with Search Tool for the Retrieval of Interacting Genes (STRING, string-db.org) and MCODE (Molecular Complex Detection) app in Cytoscape software. With prognostic information and expression data from UALCAN and Gene Expression Profiling Interactive Analysis (GEPIA) (9, 10), we identified three significant genes (*TNC, SPP1* and *KIT*). After Kyoto Encyclopedia of Genes and Genomes (KEGG) analysis, all three genes were located in the PI3K-Akt Signaling Pathway. Our results provide additional genetic biomarkers for SKCM patients.

## Material and methods

### Gene expression data

Gene expression profiles data was obtained from GEO. Microarrays of GSE31909, GSE104849 and GSE111766, which contained 13 melanoma samples and 12 melanocyte samples, were used to identify DEGs. All microarrays were performed on Platform of GPL10558 (Illumina HumanHT-12 V4.0 expression beadchip). After GEO2R analyzing, DEGs with |logFC|>2 and P value< 0.05 between melanoma samples and normal melanocyte samples were identified. DEGs with log FC< 0 was considered as down-regulated genes, and DEGs with log FC>0 was considered as up-regulated genes. Venn software was used to detect the intersection of DEGs in the three datasets.

### DEGs function and interaction analysis

Gene ontology (GO) analysis and KEGG analysis were performed with the Database for Annotation, Visualization and Integrated Discovery (DAVID, http://david.ncifcrf.gov/home.jsp), which is designed to identify genes or proteins function to understand biological meaning behind large list of genes (11, 12). STRING was used to evaluate protein-protein interaction among DEGs. MCODE app in Cytoscape software was used to reanalyze the modules of the PPI network to identify core genes.

### Survival rate analysis

UALCAN, an interactive web resource containing The Cancer Genome Atlas (TCGA) and MET500 cancer OMICS data, was used to analyze patient survival information for core genes. After survival analysis, genes with P< 0.05 were reevaluated with GEPIA to analyze the tumor/normal differential expression. Core genes with significant differential survival rate and expression were reanalyzed with DAVID to identify the key pathways related with SKCM prognosis.

## Results

### Identification of DEGs in skin cutaneous melanoma

After GEO2R analysis, we extracted 248, 401 and 246 DEGs from GSE31909, GSE104849 and GSE111766, respectively. Then, we used Venn diagram software to identify the commonly DEGs in the three datasets. Results showed that a total of 63 commonly DEGs were detected, including eight up-regulated genes (logFC>0) and 55 down-regulated genes (logFC< 0) in 13 melanoma samples and 12 melanocyte samples in our present study. (Table 1 & Fig. 1).

**Fig. 1.**
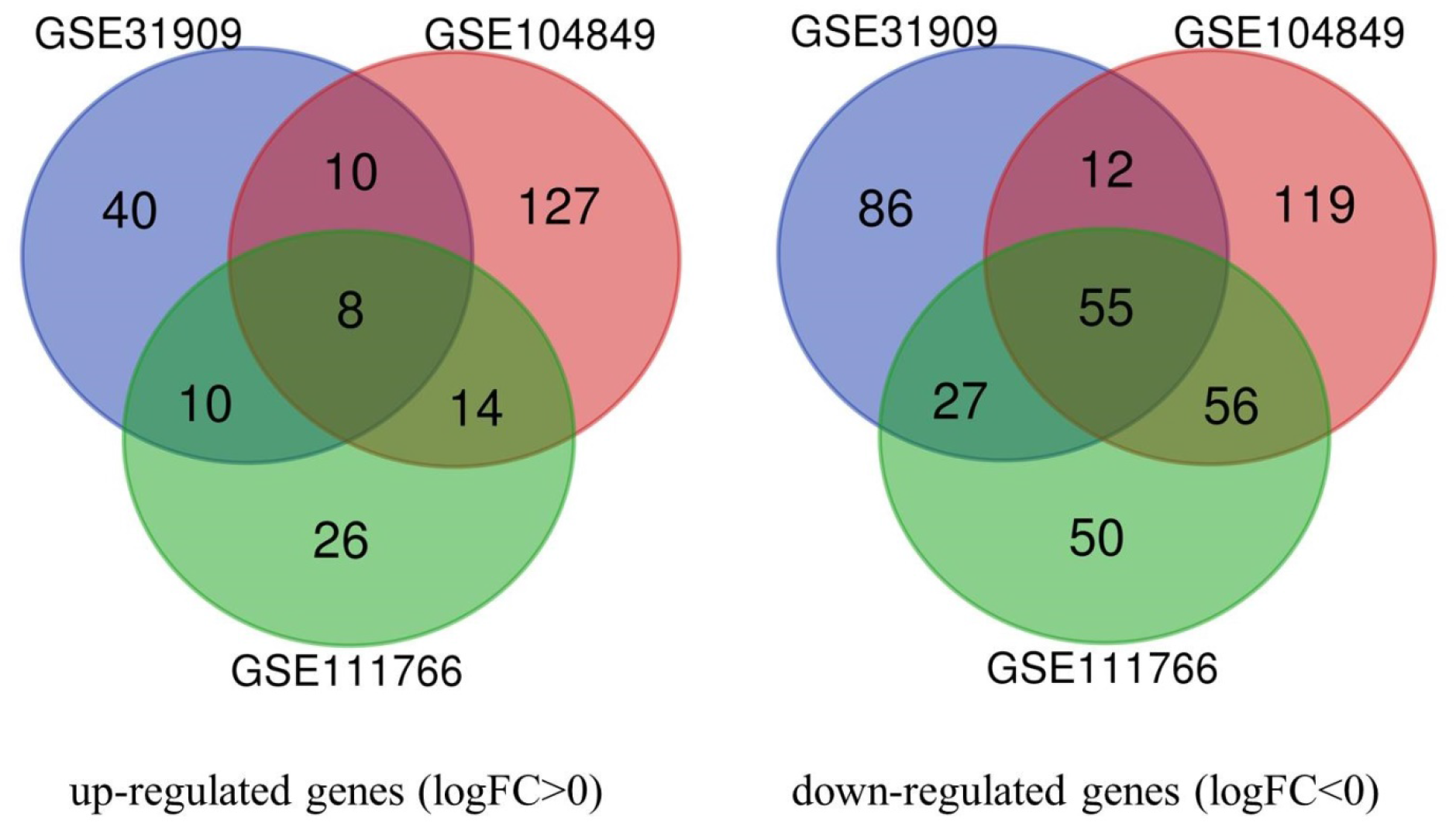
Venn diagrams of DEGs from three datasets. Eight up-regulated genes and 55 down-regulated genes were presented in all three datasets

**Table 1.**
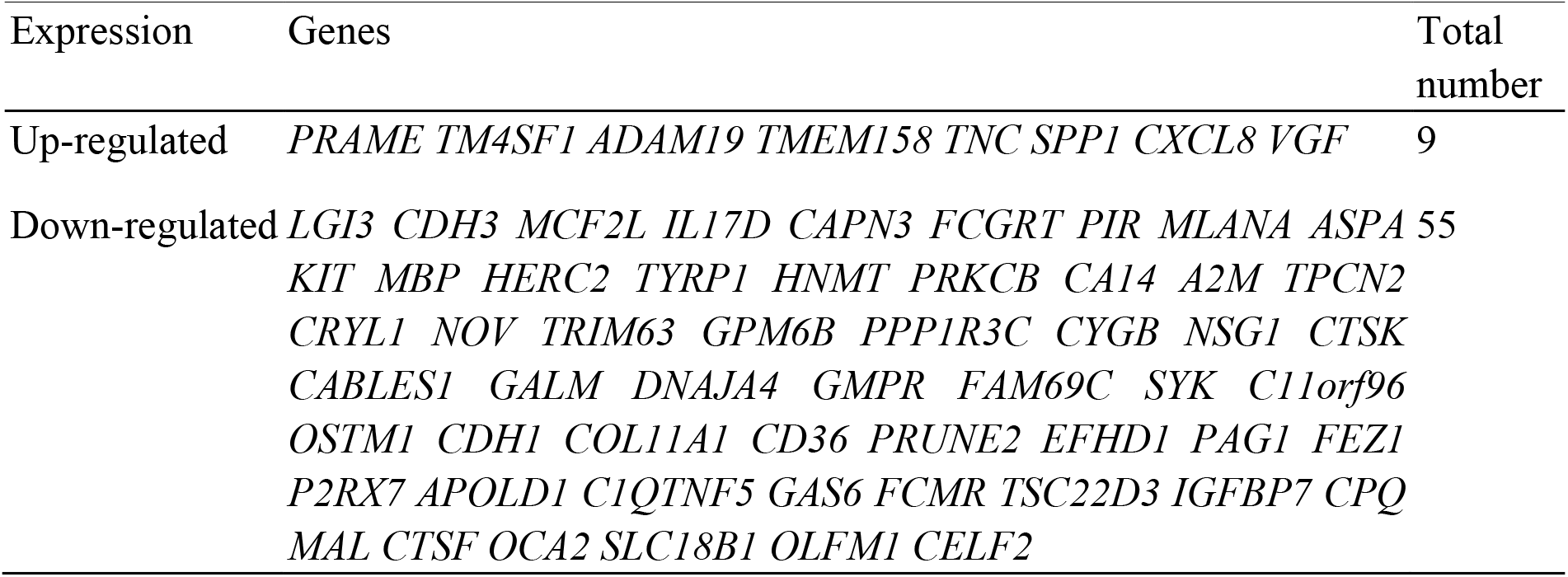
64 differentially expressed genes presented in all three datasets

### GO analysis and KEGG pathway analysis of DEGs

GO analysis and KEGG pathway analysis were performed for eight up-regulated genes and 55 down-regulated genes by DAVID software. For up-regulated genes, GO analysis results revealed that these genes were enriched in biological processes like osteoblast differentiation and extracellular matrix organization, and cell component like extracellular space. For down-regulated genes, in the category of biological processes, genes were major enriched in positive regulation of gene expression, intracellular signal transduction, cell adhesion and apoptotic process. In the category of cell component, genes were major enriched in cytoplasm, plasma membrane, cytosol and extracellular space. In the category of molecular function, genes were major enriched in receptor binding, cysteine-type peptidase activity, protease binding and lipid binding (Table 2).

**Table 2.**
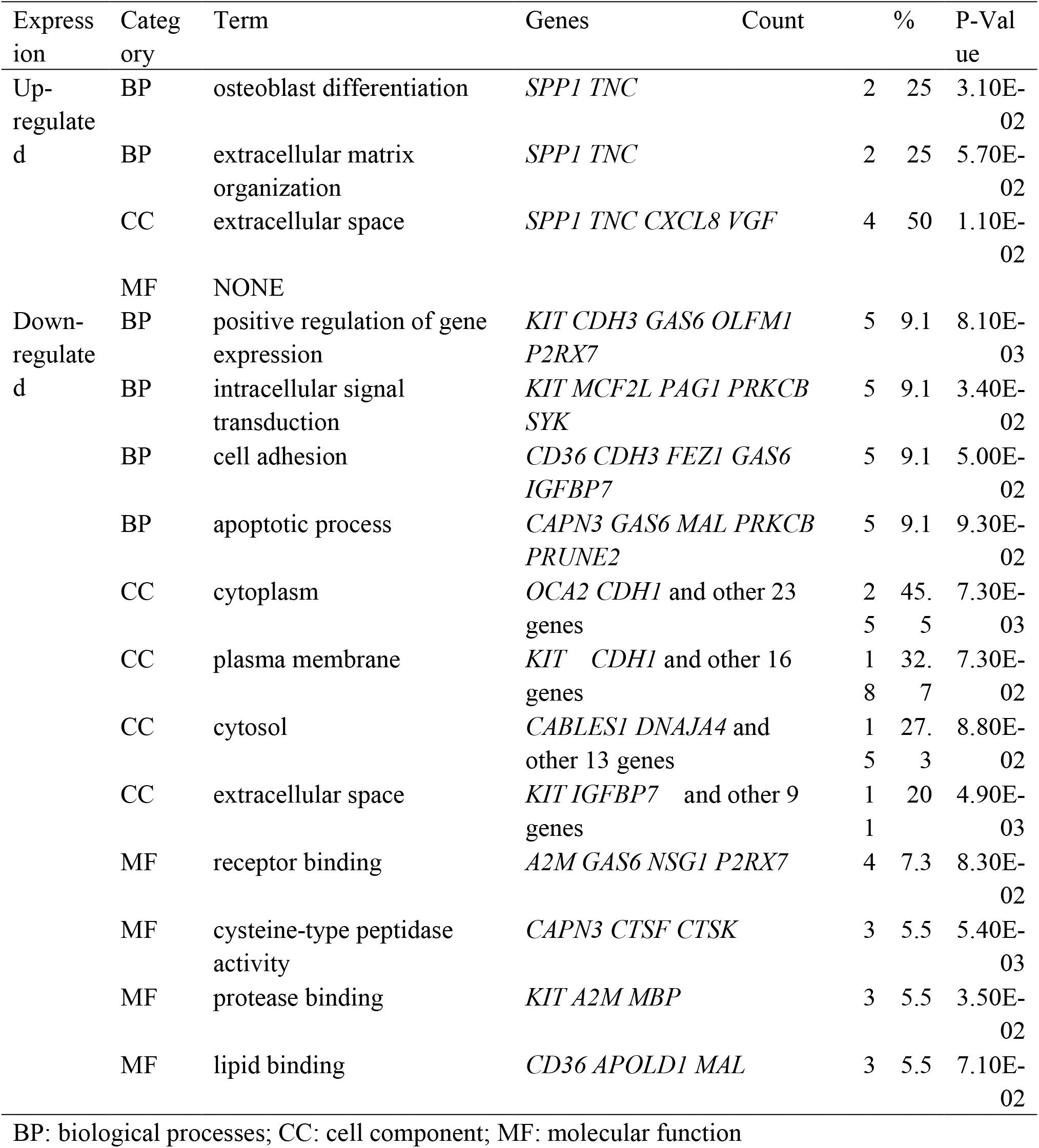
Gene ontology analysis of differentially expressed genes

KEGG analysis results demonstrated that up-regulated genes were particularly enriched in ECM-receptor interaction, toll-like receptor signaling pathway, focal adhesion and PI3K-Akt signaling pathway. Down-regulated genes were enriched in melanogenesis and histidine metabolism (Table 3).

**Table 3.**
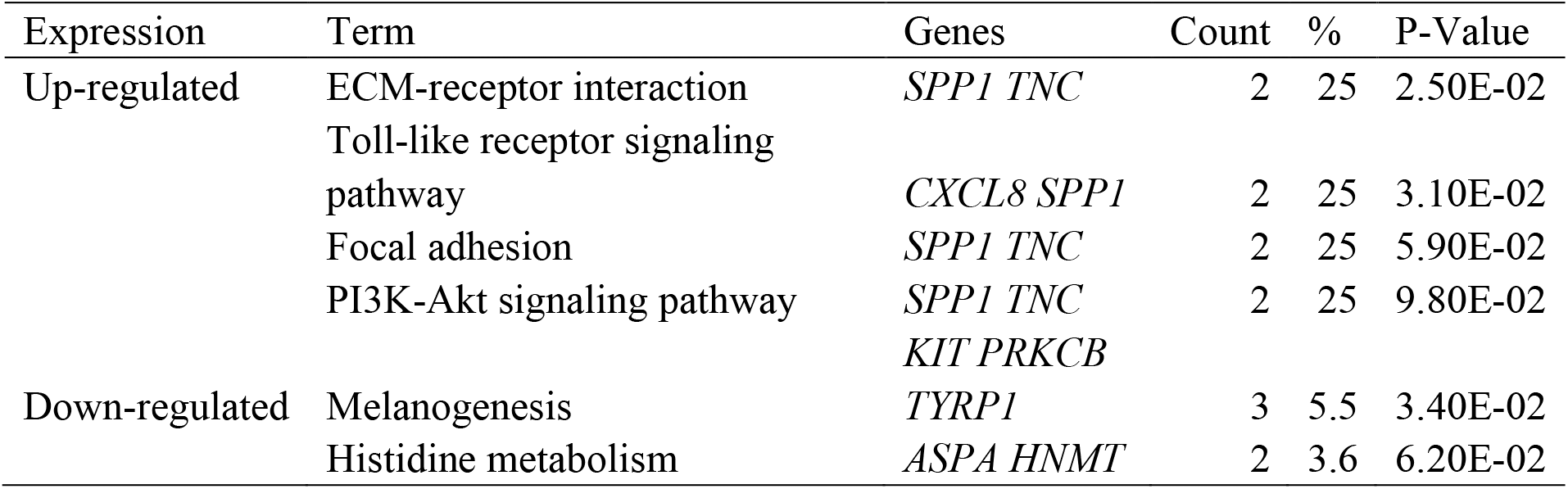
KEGG pathway analysis of differentially expressed genes

### Protein–protein interaction network (PPI) analysis

After STRING analysis of all 63 DEGs in common from three datasets, 30 of them were imported into the PPI network complex which included six up-regulated genes and 24 down-regulated genes. Cytoscape MCODE revealed two overlapped clusters (Fig. 2). 20 core genes were located in the two clusters including four up-regulated genes and 16 down-regulated genes (Fig. 2).

**Fig. 2.**
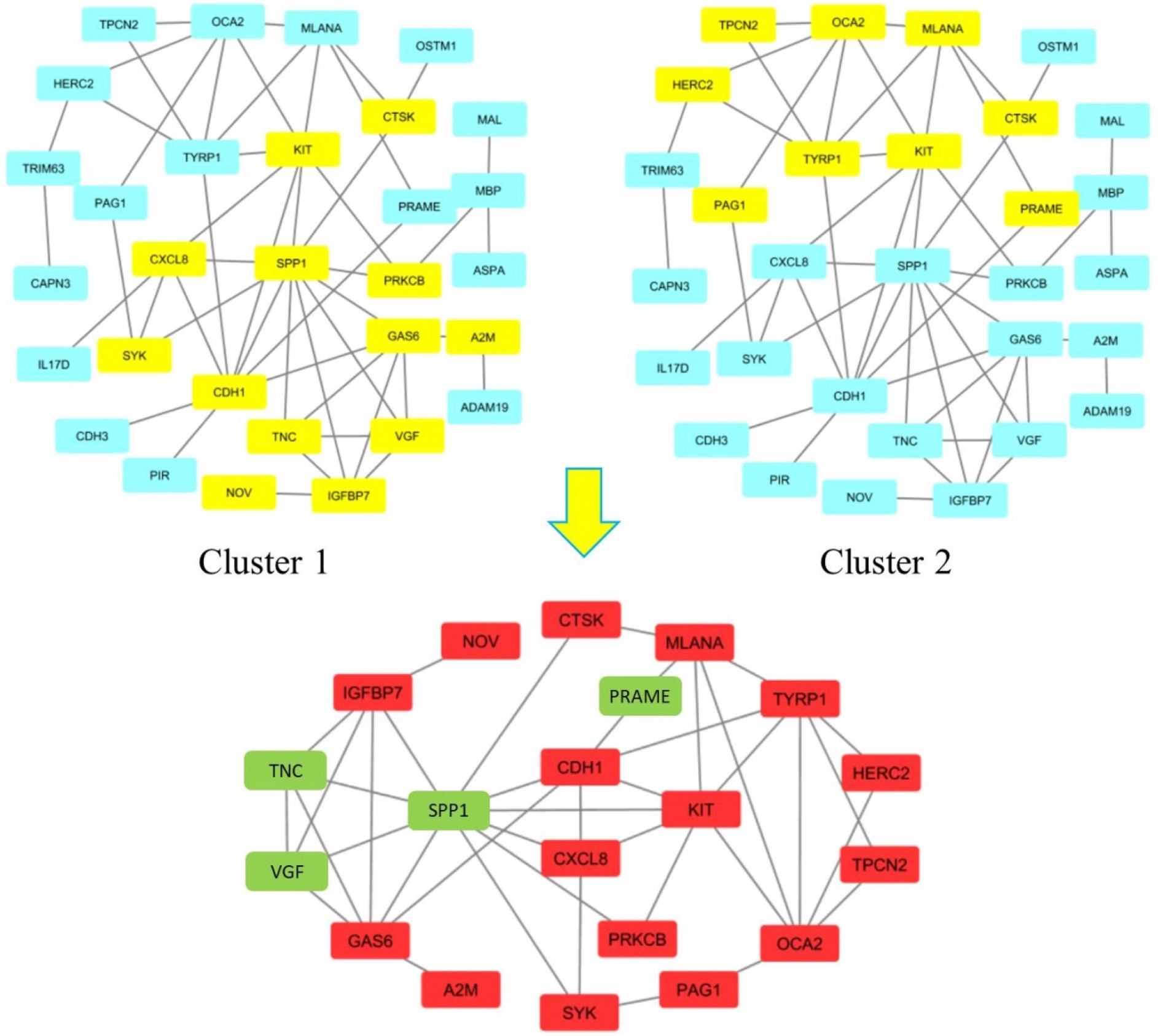
Cluster analysis of 30 DEGs by Cytoscape MCODE. Genes in yellow were located in two overlapping clusters. Genes in green were up-regulated genes. Genes in red were down-regulated genes.

### Survival rate and expression analysis

Patient survival information for core genes was analyzed by UALCAN. High expression in six genes and low expression in three genes had a significantly worse survival rate (P < 0.05, Fig. 3). GEPIA was used to reanalyze the nine gene expression level between cancerous and normal people. Results showed that three genes (*TNC, SPP1* and *KIT*) had significant expression change in melanoma samples and melanocyte samples and consisted with microarrays data (P < 0.05, Fig. 4).

**Fig. 3.**
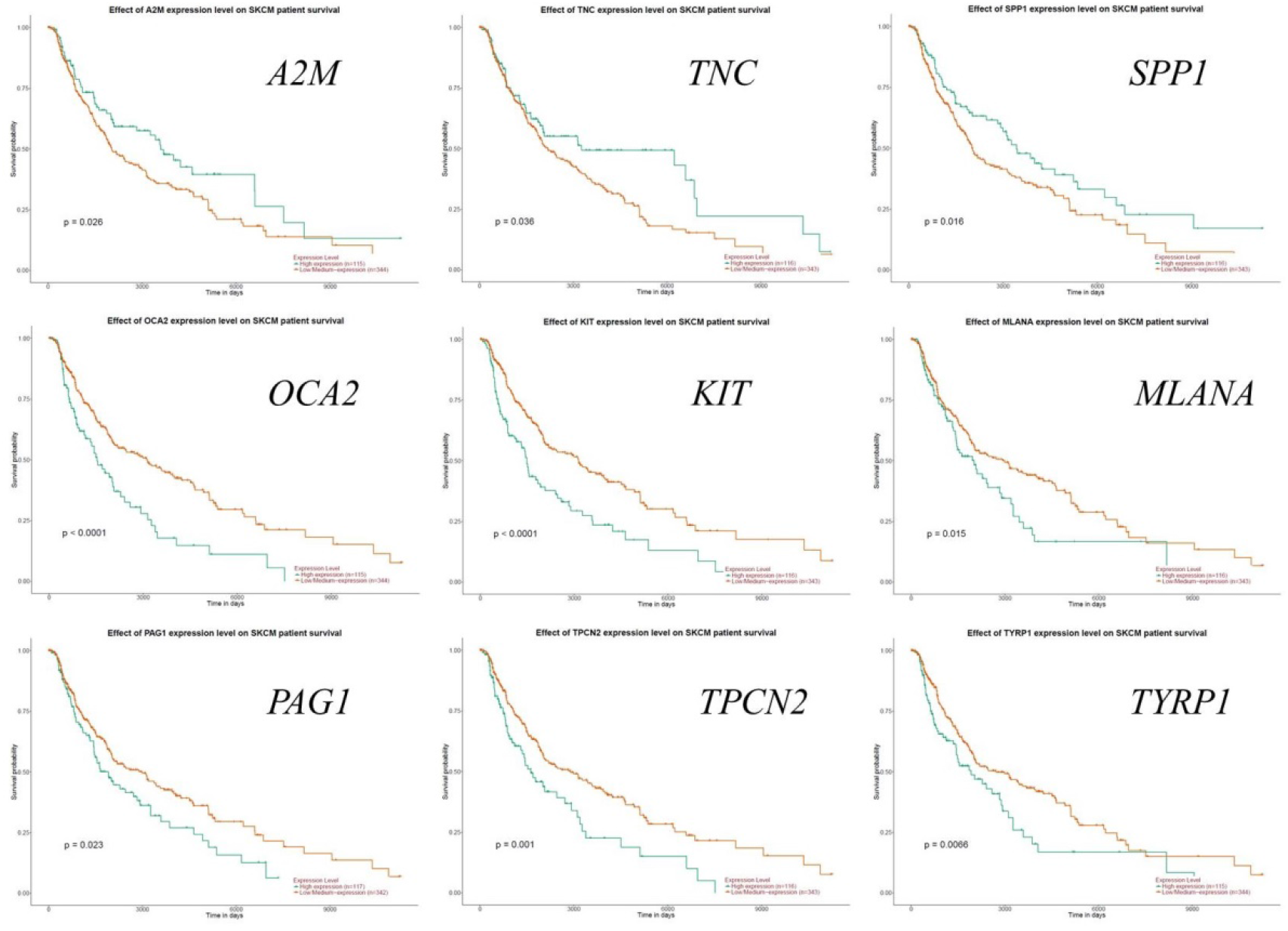
The prognostic information of the nine core genes with a significantly worse survival rate. Green line: low expression; red line: high expression (P < 0.05)

**Fig. 4.**
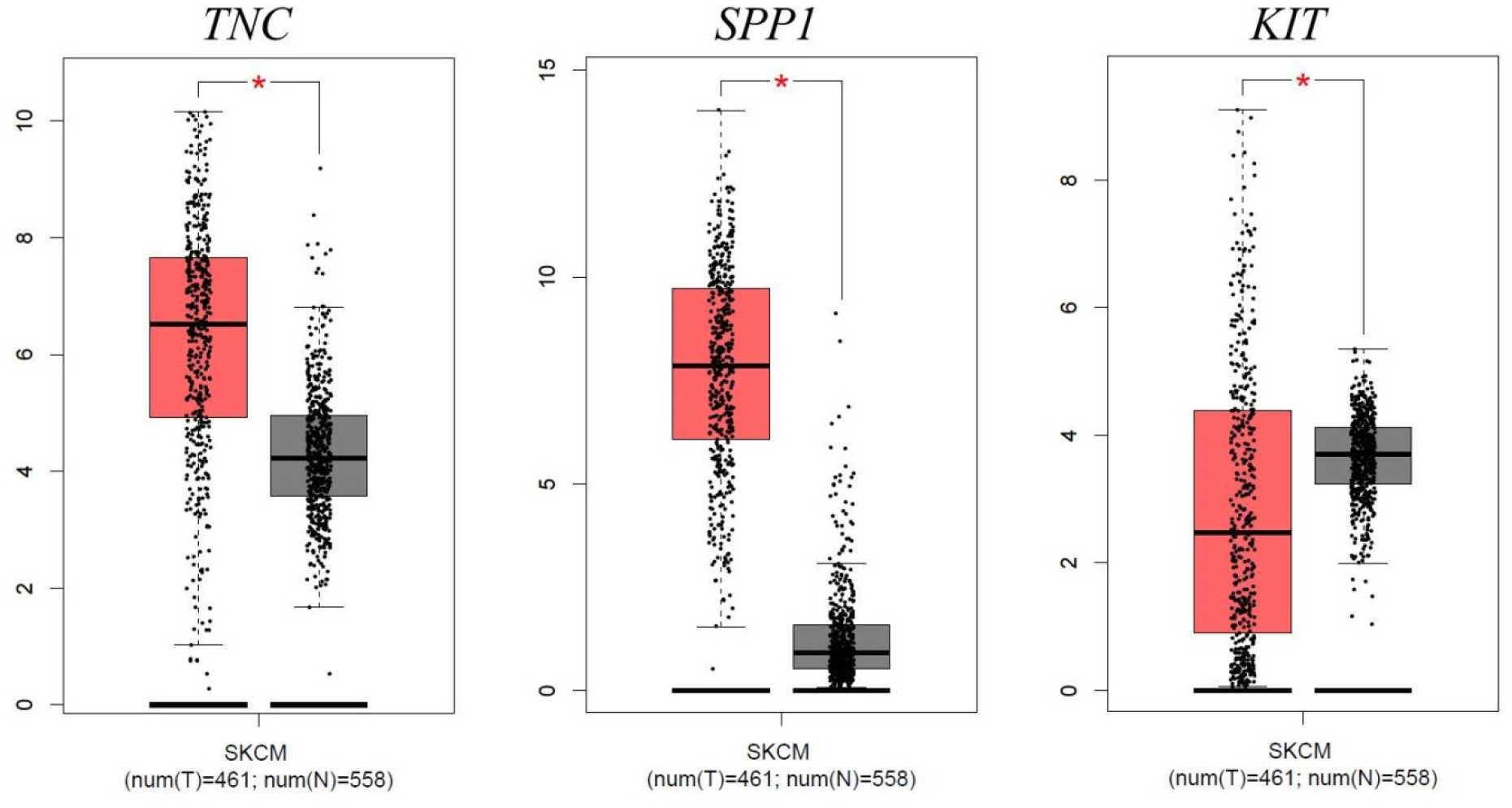
Expression level of *TNC, SPP1* and *KIT* between cancerous and normal people. Left box means tumor tissues (n = 461) and right box means normal tissues (n = 558) (P < 0.05)

### Reanalysis of three key genes via KEGG pathway enrichment

To understand the possible pathway of the three key DEGs, KEGG pathway enrichment was reanalyzed by DAVID. Results showed that all three genes markedly enriched in the PI3K-Akt signaling pathway (P = 2.5E-3, Fig. 5), while *TNC* and *SPP1* were also enriched in ECM-receptor interaction (P = 2.5E-2) and Focal adhesion (P = 5.9E-2).

**Fig. 5.**
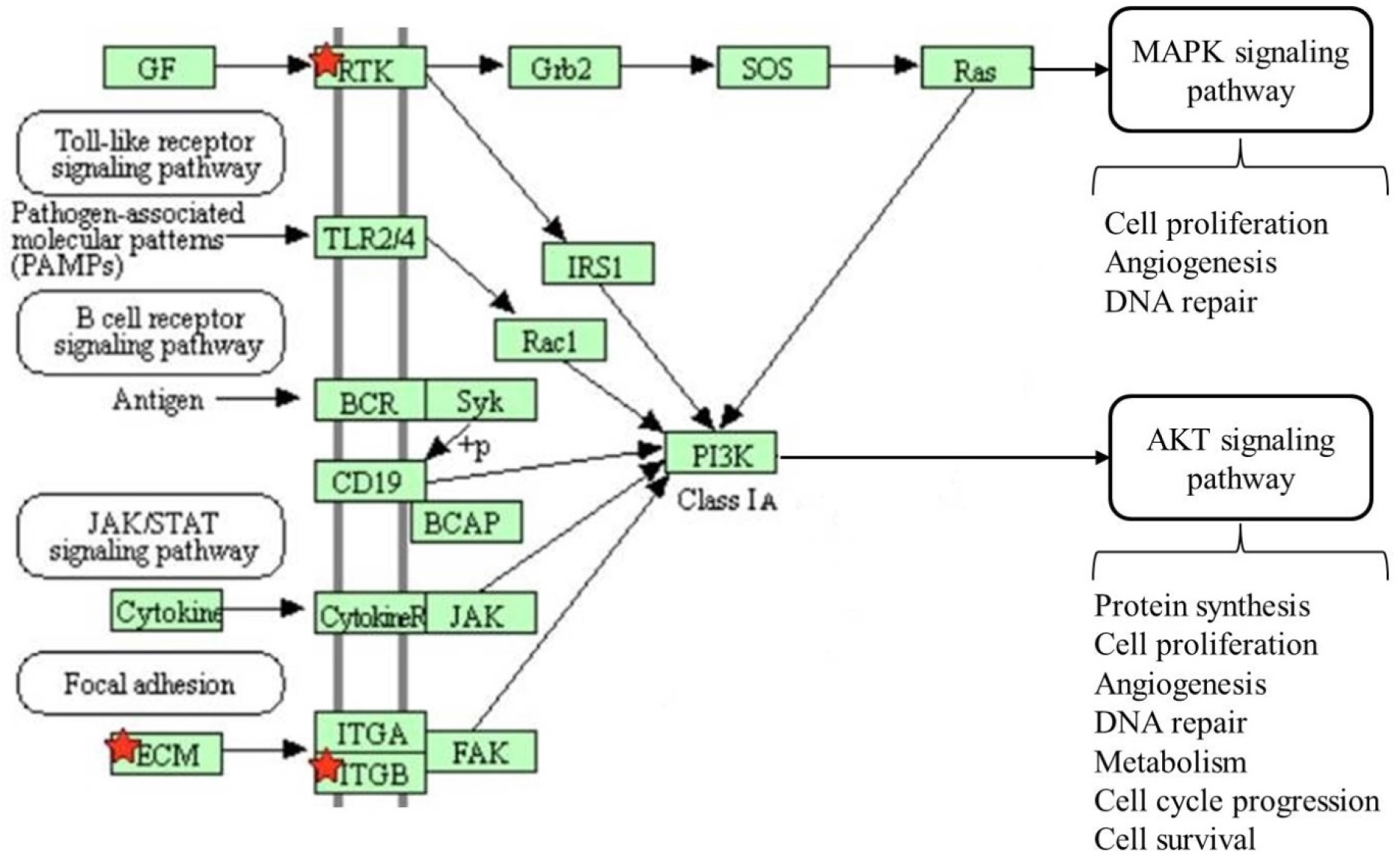
KEGG pathway reanalysis of three key genes revealed enrichment in PI3K-Akt signaling pathway.

## Discussion

SKCM is one of the most fatal skin cancers due to its high proliferative and metastatic potential. Several biomarkers for the TNM (Topography, Lymph Node and Metastasis) classification have been identified, but these staging markers often fail to predict the patient’s prognosis accurately (13). In our study, we uncovered 63 common DEGs from microarrays of GSE31909, GSE104849 and GSE111766. Protein-protein interaction analysis identified 20 core genes in the two clusters. With prognostic information and expression information in public databases, we identified three significant genes *TNC, SPP1* and *KIT*. These three genes are located in the PI3K-Akt signaling pathway, which is one of the most important signaling networks in cancer and the activation of this pathway plays a significant role in melanoma(14). PI3K-AKT pathway also plays important roles in melanoma therapeutic resistance. Therefore, our results provide further genetic markers for SKCM patients’ prognosis.

*TNC* encodes tenascin, an extracellular matrix protein with a spatially and temporally restricted tissue distribution. This protein contains multiple EGF-like and fibronectin type-III domains. The EGF-like domains is considered anti-adhesive and the fibronectin type-III domains are considered adhesive (15–17). Cell interaction with tenascin can promote migration in a mesenchymal state or an amoeboid state to penetrate denser matrix (18–20). Recent evidence also showed The EGF-like domains of TNC acted as an ultralow affinity ligand for the EGF receptor (21, 22) and the restriction of EGFR signaling increased cell survival protect stem cells from apoptosis (23, 24). Therefore, the function of highly expressed *TNC* in melanoma includes both promoting migration and invasion through the dermis, and colonization the distant target organ with supporting melanoma cells survival. The *TNC* gene expression is strictly regulated and it is lowly expressed in adult skin (25). *TNC* expression is dramatically elevated level in patients with higher stages melanoma than lower stages tumor (26). Thus, *TNC* was considered as a potential biomarker of melanoma aggressiveness and treatment effect. However, the mechanism behind the up-regulation of *TNC* remains to be further determined.

*SPP1* encodes secreted phosphoprotein 1 (also unknown as osteopontin, OPN). This protein is a matricellular protein that is produced by multiple tissues and mostly abundant in bone. It is involved in bone mineralization by attachment of osteoclasts to the mineralized bone matrix (27). SPP1 protein is produced by cancer cells and plays crucial roles in cancer progression by facilitating adhesion, migration and survival of tumor cells (28–30). Several studies indicate that SPP1 protein is overexpressed in the majority of human cancers and the up-regulated expression is related with associated with poor clinical outcome in a variety of cancers including breast cancer, lung cancer, liver cancers, prostate cancer, etc. (31–33). Similar to *TNC, SPP1* overexpression was significantly associated with tumor thickness, metastasis, and reduced relapse-free survival in melanoma patients (34, 35). Therefore, *SPP1* is a promising biomarker for detecting melanoma aggressiveness and a factor for the survival prediction in patients.

*KIT* encodes Mast/Stem Cell Growth Factor Receptor. It is the human homolog of the proto-oncogene c-kit. KIT protein is a Receptor Tyrosine Kinase containing a extracellular ligand-binding domain, a transmembrane domain, an juxta-membrane domain and two tyrosine kinase domains (36). *KIT* loss-of-function mutations result in piebaldism, which is characterized by the congenital absence of melanocytes in affected areas of the skin and hair (37, 38). Somatic *KIT* mutations that result in KIT receptor tyrosine kinase constitutive activity, were reported to cause a variety of cancers like gastrointestinal stromal tumor, acute myelogenous leukemia, testicular germ cell tumor (39, 40), as well as melanoma (mostly in mucosal and acral melanomas) (41–43). However, activating mutations in the *KIT* gene do not necessarily correlate with c-KIT expression (44, 45). *KIT* has been considered a melanoma suppressor and the expression appears to be downregulated or lost in invasive and metastatic melanomas (46, 47). Loss of *KIT* expression were also seen in other cancer (48, 49) and may be a required step in tumor progression due to its important roles in cell differentiation and tissue morphogenesis. Although *KIT* expression is down-regulated in most subsets of melanomas (including SKCM), one study showed human posterior uveal melanoma expresses *KIT* at high levels frequently (50). This result indicated that the *KIT* expression in melanomagenesis is more complex and need further investigation.

We identified three potential predictors (*TNC, SPP1* and *KIT*) for survival rates of patients with SKCM based on three different microarray datasets. These three genes are located in the PI3K-Akt signaling pathway and were reported to have important roles in the progression of SKCM. Further researches are needed to verify these prognosis bio-markers and improve the clinical outcomes of patients with this highly aggressive cancer.

## Data Availability

The datasets generated and/or analyzed during the current study are available in the GEO repository (https://www.ncbi.nlm.nih.gov/geo/query/acc.cgi?acc=GSE31909), (https://www.ncbi.nlm.nih.gov/geo/query/acc.cgi?acc=GSE104849) and (https://www.ncbi.nlm.nih.gov/geo/query/acc.cgi?acc=GSE111766).

https://www.ncbi.nlm.nih.gov/geo/query/acc.cgi?acc=GSE104849

https://www.ncbi.nlm.nih.gov/geo/query/acc.cgi?acc=GSE111766

## Acknowledgements

This study was partially supported by Scientific Research Cultivation Fund of Capital Medical University (PYZ19034).

## Availability of data and materials

The datasets generated and/or analyzed during the current study are available in the GEO repository (https://www.ncbi.nlm.nih.gov/geo/query/acc.cgi?acc=GSE31909),(https://www.ncbi.nlm.nih.gov/geo/query/acc.cgi?acc=GSE104849) and (https://www.ncbi.nlm.nih.gov/geo/query/acc.cgi?acc=GSE111766).

## Ethics approval and consent to participate

Not applicable.

